# Machine Unlearning for GDPR Right-to-Erasure in Antimicrobial Resistance Prediction Models

**DOI:** 10.64898/2026.03.09.26347960

**Authors:** Saniya, Abdullah Ahmad Khan

## Abstract

**Objective:** Healthcare machine learning models trained on patient data must comply with the General Data Protection Regulation (GDPR) right-to-erasure requirement, which mandates the removal of individual data contributions from deployed models. Full retraining, the current standard, is computationally expensive. This study evaluates Sharded, Isolated, Sliced and Aggregated (SISA) training as an efficient framework for predicting antimicrobial resistance (AMR).

**Materials and Methods:** SISA training (5 shards) was compared with Full Retraining, Label-Flip Retraining, Influence Reweighting, and Selective Tree Pruning on two datasets: the Antibiotic Resistance Microbiology Dataset (ARMD; *n* = 1,245,767 EHR records) and the BV-BRC/PATRIC genomic surveillance dataset (*n* = 400,372). Random Forest classifiers used 500 estimators. Metrics included accuracy, AUC-ROC, membership inference attack (MIA) gap, unlearning time, and cumulative 12-month deletion cost.

**Results:** SISA achieved an 8.9× speedup over full retraining on ARMD (7.5 s vs. 66.7 s) and a 9.8 × speedup on PATRIC (1.4 s vs. 13.4 s), with accuracy costs of 0.024% and 0.048%, respectively, both below the 0.5% clinical threshold. Label-Flip Retraining and Influence Reweighting provided no speedup (≤ 1.0 ×), while Tree Pruning exceeded the threshold on EHR data (+0.648%). Over 12 months at 50 monthly deletions, SISA reduced cumulative overhead from 800 s to 90 s (ARMD) and from 160 s to 16 s (PATRIC).

**Discussion:** SISA maintains predictive performance while reducing computational cost, supporting machine unlearning for regulatory compliance in clinical ML systems.

**Conclusion:** SISA provides an efficient framework for maintaining GDPR-compliant AMR prediction models and support the scalable processing of patients deletion requests.

## 1. INTRODUCTION

Antimicrobial resistance (AMR) is a critical global health threat, with resistant infections causing an estimated 1.27 million attributable deaths in 2019 and projected to become a leading cause of mortality by 2050. [1, 2] Machine learning (ML) models trained on electronic health records (EHR) and genomic surveillance data have demonstrated strong potential for predicting resistance phenotypes and informing antimicrobial stewardship programmes. [3–6]

A critical and underappreciated regulatory challenge accompanies clinical ML deployment. GDPR Article 17 Right to Erasure requires that patient data be removable not only from storage but also from any trained model upon patient request. [7] Equivalent provisions exist in the Australian Privacy Act 1988 and the California Consumer Privacy Act. Full model retraining, the most straightforward compliance mechanism, is computationally prohibitive. A model trained on over one million records requires ∼67 s per deletion request, accumulating to >800 s annually at 50 monthly requests.

Machine unlearning [8] seeks computationally efficient alternatives to full retraining. The Sharded, Isolated, Sliced, and Aggregated (SISA) framework [9] partitions training data into independent shards, limiting each deletion to retraining only the affected shard. Prior SISA evaluations have focused on image classification benchmarks and have not addressed the specific characteristics of clinical AMR prediction: heterogeneous feature spaces, imbalanced phenotype distributions, strict accuracy requirements, and the need for generalization across EHR and genomic data modalities. This study provides the first systematic comparison of machine unlearning methods for AMR prediction across two independent datasets.

## 2. BACKGROUND AND SIGNIFICANCE

### 2.1 Machine unlearning

Cao and Yang [8] formally defined machine unlearning as causing a trained model to behave as if specified training data had never been observed. Bourtoule et al. [9] introduced SISA training, in which non-overlapping shards and independent sub-models reduce the deletion cost to approximately 1*/k* of full retraining when *k* shards are used.

Alternative approaches to machine unlearning have also been proposed, including gradient-based forgetting [10], amnesiac methods that reverse stored parameter updates [11], influence function approximations [12], and recent theoretical frameworks for efficient unlearning [13]. Earlier work also explored formal approaches to removing individual data contributions from trained models [14]. Certified data removal techniques have additionally been proposed to guarantee that deleted data no longer influences model predictions [15]. To date, however, these approaches have not been evaluated for clinical AMR prediction.

### 2.2 Privacy verification

Membership Inference Attacks (MIA) [16] assess whether an adversary can identify training records from model outputs. We quantify unlearning completeness using the MIA gap:

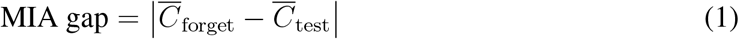

where 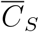 is the mean prediction confidence on set *S*. Smaller post-unlearning gaps indicate more complete erasure.

### 2.3 Significance

AMR prediction models trained on patient EHR data are directly subject to right-to-erasure provisions, yet no published study has evaluated unlearning strategies for this class of models. This study addresses that gap, providing evidence-based guidance for clinical informatics practitioners designing GDPR-compliant AMR decision-support systems [17, 18].

## 3. MATERIALS AND METHODS

### 3.1 Datasets

*ARMD*. The Antibiotic Resistance Microbiology Dataset [4] contains de-identified microbiological culture and susceptibility records from two Stanford Health Care hospitals (1999–2024). After merging the cohort, demographics, and ward tables and filtering to binary Resistant/Susceptible phenotypes, the processed dataset comprised *n* = 1,245,767 records. Features include organism type, antibiotic agent, patient age, gender, and ward classification. The resistance rate was ≈37%. The dataset is available under a Data Use Agreement (DUA); registration required at https://clinicalml.org/data/amr-dataset.

*PATRIC/BV-BRC*. The NIH Bacterial and Viral Bioinformatics Resource Center [19] provides genomic AMR phenotype data from global published studies and GenBank. Records were retrieved via the BV-BRC REST API, querying Resistant and Susceptible phenotypes separately (up to 250,000 per class) to ensure class balance, yielding *n* = 400,372 records. Features include bacterial species, antibiotic agent, and log-transformed minimum inhibitory concentration (MIC), laboratory method, and host. Dataset characteristics are summarised in Table 1.

**Table 1.** Summary of the two AMR datasets used in this study.

| Characteristic | ARMD | PATRIC/BV-BRC |
| --- | --- | --- |
| Data modality | Clinical EHR | Genomic surveillance |
| Source | Stanford Health Care,<br>1999–2024 | NIH BV-BRC / GenBank |
| Total records | 1,245,767 | 400,372 |
| Forget set | 500 patients | 500 genomes |
| Train/Test split | 80%/20% stratified | 80%/20% stratified |
| Resistance rate | $\approx 37\%$ | $\approx 62.5\%$ |
| Key features | Organism, antibiotic, age,<br>gender, ward | Species, antibiotic, MIC,<br>method, host |

### 3.2 Ethical considerations

This study used only de-identified datasets and did not involve direct interaction with human participants. The ARMD dataset was accessed under the MIT ClinicalML Data Use Agreement, which permits secondary analysis of de-identified clinical records; no additional institutional ethics approval was required. The PATRIC/BV-BRC dataset is an open-access genomic surveillance resource with no human subject considerations.

### 3.3 Experimental design

A forget set of 500 unique records (random seed 42) was designated per dataset, representing a realistic monthly GDPR deletion batch. The remaining data were split into a retain (80%) and a test (20%) set using stratified sampling. All experiments used a Random Forest classifier [20] with 500 estimators, maximum depth 12, minimum 5 samples per leaf, and `n_jobs=-1`. Categorical features were one-hot encoded, retaining the top 20–30 categories per feature. MIC values were log-transformed as *x*^*′*^ = log(1 + *x*). Wall-clock unlearning time was measured using Python’s `time` module, excluding original model training time. Because both datasets contain hundreds of thousands to over one million records, performance variance across random seeds is expected to be minimal; therefore, a fixed seed was used to ensure deterministic reproducibility.

### 3.4 Ethics and Data Governance

This study used only de-identified datasets and did not involve direct interaction with human participants. The ARMD dataset consists of anonymized electronic health record data accessed under the MIT ClinicalML Data Use Agreement, which permits secondary analysis of de-identified clinical records for research purposes. The BV-BRC/PATRIC dataset contains publicly available genomic surveillance data.

Because all data were fully de-identified and no identifiable patient information was Information was accessed, and the study did not require additional institutional review board (IRB) approval in accordance with standard guidelines for secondary analysis of anonymized data.

### 3.5 Unlearning methods

*Full Retraining (Gold Standard)*. A new Random Forest is trained from scratch on the retained set. Provides exact unlearning at maximum computational cost (*T*_retrain_); all speedup values are reported relative to this baseline.

*SISA Training*. Following Bourtoule et al., [9] the retain set is partitioned into *k* = 5 equal shards. Five sub-models *{f*_1_, …, *f*_5_*}* predict by averaging class probabilities:

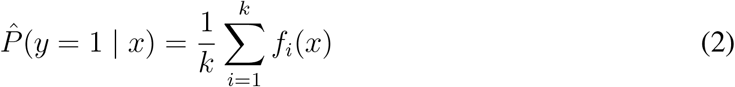

Deletions retrain only the affected shard; theoretical cost ≈ *T*_retrain_*/k*.In the present experiments, the designated forget set was assumed to lie within a single shard, representing a best-case deletion scenario and yielding the maximum theoretical speedup. In real-world deployments, patient deletion requests may span multiple shards, thereby increasing retraining costs in proportion.

#### Label-Flipped Retraining

Forget-set records are relabelled to the opposite class (0 ↔ 1), and the model is retrained on retain *∪* relabelled-forget. This heuristic baseline is inspired by gradient-ascent forgetting approaches proposed for neural networks [10], but implemented here as label-flipped retraining to produce an adversarial signal for tree-based models that lack differentiable parameters.

#### Influence Reweighting

Based on influence function theory, [12] forget set records receive sample weight *w* = 10^−6^ during full-dataset retraining.

#### Selective Tree Pruning

Trees with an error rate below the 40th percentile on the forget set are removed:

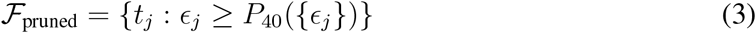

No retraining is required, yielding the fastest method.

### 3.6 Evaluation metrics

Performance was assessed by accuracy and AUC-ROC on the held-out test set. A conservative tolerance of ≤0.5% relative accuracy degradation from the original model was used as an operational threshold for clinical decision support systems. In large clinical datasets with stable baselines, accuracy variations below this level are typically within routine retraining variance and unlikely to affect treatment decisions; methods exceeding this margin were therefore considered to violate acceptable performance drift. Privacy was assessed using the MIA gap (Equation 1). Efficiency metrics included wall-clock unlearning time, speedup *S* = *T*_retrain_*/T*_method_, and cumulative 12-month deletion cost *C*_12_ = 12 × *T*_method_ at 50 requests/month.

## 4. RESULTS

### 4.1 ARMD clinical EHR dataset

Table 2 presents results on ARMD (*n* = 1,245,767; 500 trees; 500 forget patients). The original model achieved 84.89% accuracy and AUC-ROC 0.808 (MIA gap 3.359 × 10^−3^; training time 66.7 s).

**Table 2.** Machine unlearning method comparison on ARMD (*n* = 1,245,767 records; 500 trees; forget set = 500 patients). ^*∗*^Training time (not unlearning time). ^*†*^Violates the 0.5% clinical accuracy threshold. Bold row indicates the proposed SISA method. ↓ lower is better; ↑ higher is better.

| Method | Acc.↑ | AUC↑ | —MIA—<br>$\times 10^{-3}$ ↓ | Time<br>(s)↓ | Speedup↑ | Acc.<br>Drop↓ |
| --- | --- | --- | --- | --- | --- | --- |
| Original | 84.89% | 0.808 | 3.359 | 66.7* | — | — |
| Full Retrain | 84.90% | 0.807 | 3.207 | 66.7 | 1.0× | −0.012% |
| **SISA** | **84.91%** | **0.808** | **3.316** | **7.5** | **8.9×** | **−0.024%** |
| Label-Flip<br>Retraining | 84.89% | 0.807 | 2.591 | 81.6 | 0.8× | −0.006% |
| Influence Rewt. | 84.89% | 0.807 | 3.269 | 80.1 | 0.8× | −0.004% |
| Tree Pruning | 84.24% | 0.801 | 3.403 | 0.8 | 78.5× | +0.648%<sup>†</sup> |

SISA achieved an **8.9**× **speedup** (7.5 s vs. 66.7 s) with a −0.024% accuracy cost, well within the 0.5% threshold. Label-Flip Retraining and Influence Reweighting were *slower* than full retraining (0.8×) because both require training on the full dataset. Tree Pruning was fastest (78.5×) but violated the clinical threshold (+0.648%), disqualifying it from EHR deployment. SISA accuracy is marginally higher than the original model (≤ 0.05%) due to the stochastic nature of Random Forest training (bootstrap sampling and feature subsampling); removing a small number of records can slightly alter the tree structure and occasionally improve generalization. The magnitude of this difference is extremely small (*<* 0.1%) and should be interpreted as statistical noise rather than a systematic performance improvement.

### 4.2 PATRIC/BV-BRC genomic dataset

Table 3 presents results on PATRIC (*n* = 400,372; 500 trees; 500 forget genomes). The original model achieved 67.01% accuracy and AUC-ROC 0.722. The lower baseline relative to ARMD reflects the greater phenotypic heterogeneity of global genomic surveillance data.

**Table 3.** Machine unlearning method comparison on PATRIC/BV-BRC (*n* = 400,372 records; 500 trees; forget set = 500 genomes). Notation as per Table 2.

| Method | Acc.↑ | AUC↑ | —MIA—<br>$\times 10^{-3} \downarrow$ | Time<br>(s)↓ | Speedup↑ | Acc.<br>Drop↓ |
| --- | --- | --- | --- | --- | --- | --- |
| Original | 67.01% | 0.722 | 4.733 | 13.4* | — | — |
| Full Retrain | 67.03% | 0.720 | 4.593 | 13.4 | 1.0× | −0.015% |
| **SISA** | **67.06%** | **0.721** | **4.870** | **1.4** | **9.8×** | **−0.048%** |
| Label-Flip<br>Retraining | 66.62% | 0.716 | 4.030 | 13.3 | 1.0× | +0.389% |
| Influence Rewt. | 67.03% | 0.720 | 4.656 | 13.7 | 1.0× | −0.016% |
| Tree Pruning | 67.06% | 0.719 | 3.226 | 0.2 | 70.4× | −0.045% |

SISA again demonstrated the strongest profile: **9.8**× **speedup** with a −0.048% accuracy cost. Label-Flip Retraining degraded accuracy by 0.389%, approaching the clinical threshold. Tree Pruning was within threshold on PATRIC (−0.045%) but, given its violation on ARMD, cannot be recommended as a cross-modality solution.

### 4.3 Cross-dataset consistency

Figure 1 presents the cross-dataset comparison. SISA is the only evaluated method to achieve consistent speedups (≥8.9×) with an accuracy cost below 0.5% on both datasets under the experimental shard configuration. Label-Flip Retraining and Influence Reweighting provide no improvement on either dataset. Tree Pruning’s variable accuracy cost (0.648% EHR vs. 0.045% genomic) demonstrates modality-specific sensitivity that precludes its general-purpose use.

**Figure 1.**
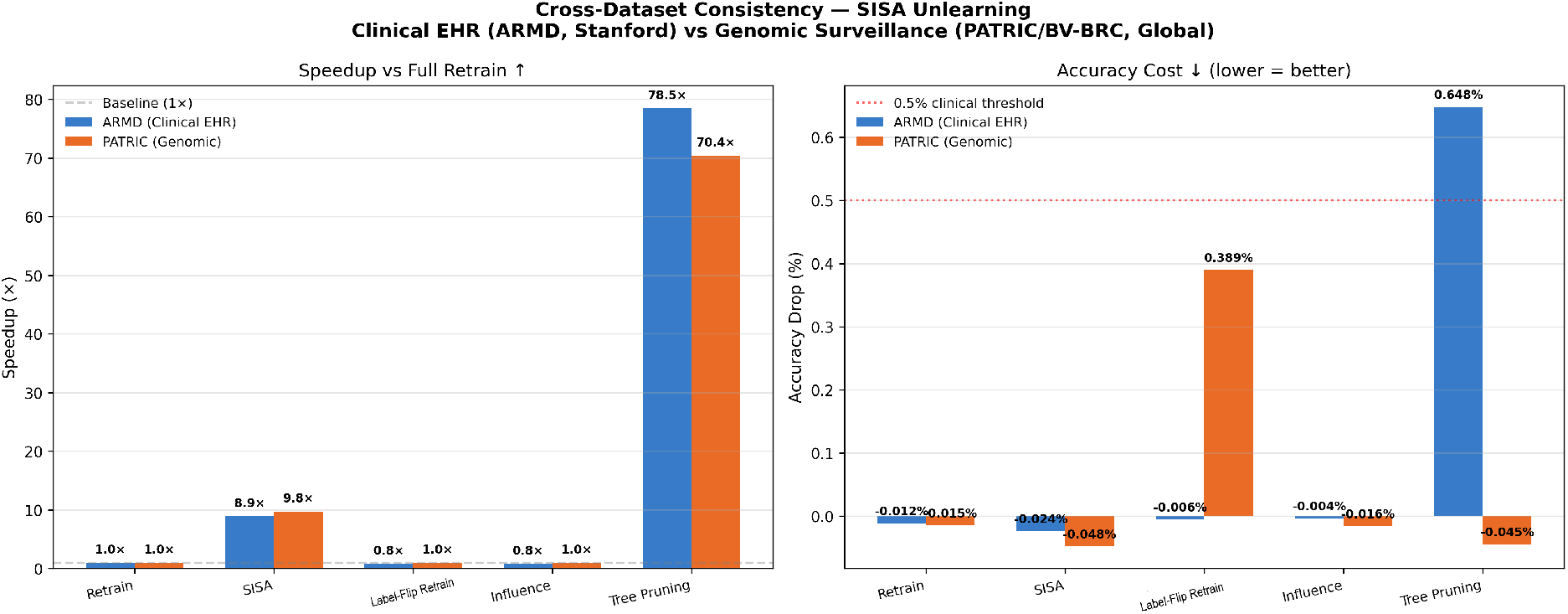
Cross-dataset consistency of unlearning methods. **Left**: Speedup relative to full retraining on ARMD (clinical EHR, blue bars) and PATRIC (genomic, orange bars). SISA achieves consistent 8.9–9.8× speedup across both datasets; Label-Flip Retraining and Influence Reweighting provide no improvement (≤1×). **Right**: Accuracy cost of each unlearning method. The red dotted line marks the 0.5% clinical threshold. Tree Pruning violates this threshold on EHR data (+0.648%), while SISA remains within bounds on both datasets. Alt text: Two grouped bar charts. The left chart shows speedup values for five unlearning methods on two datasets; SISA bars reach approximately 9 times higher than Label-Flip Retraining and Influence bars, which barely exceed the baseline. The right chart shows the accuracy drop values; a red dotted line at 0.5% marks the clinical threshold; the Tree Pruning bar for ARMD exceeds this line while all SISA bars remain below it.

### 4.4 PATRIC method comparison

Figure 2 presents the four-panel method comparison on the PATRIC dataset. SISA’s 1.4 s unlearning time (9.8× speedup) is visually distinctive while accuracy (67.06%) is maintained above all competitors.

**Figure 2.**
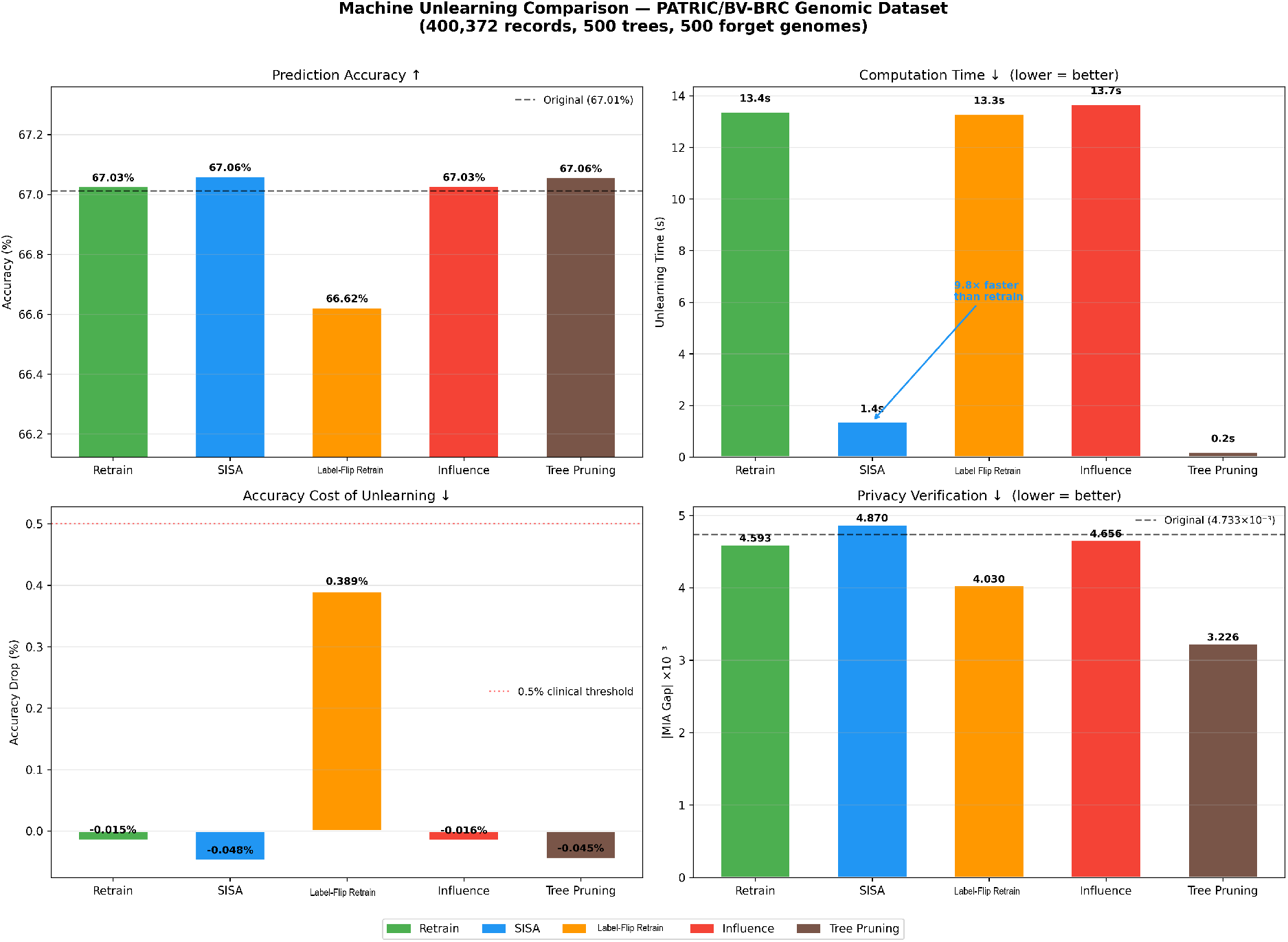
Machine unlearning method comparison on the PATRIC/BV-BRC genomic surveillance dataset (*n* = 400,372 records; 500 trees; forget set = 500 genomes). **Top-left**: Prediction accuracy after unlearning; the dashed line indicates original model accuracy (67.01%). **Top-right**: Unlearning computation time in seconds; SISA achieves a 9.8× speedup (1.4 s vs. 13.4 s for full retraining). **Bottom-left**: Accuracy cost of each method; the red dotted line marks the 0.5% clinical threshold. **Bottom-right**: Absolute MIA gap (×10^−3^) for privacy verification; the dashed line shows the original model baseline (4.733 × 10^−3^). Alt text: Four-panel bar chart comparing five machine unlearning methods on a genomic AMR dataset. Top-left panel shows prediction accuracy near 67%, with Label-Flip Retraining notably lower. Top-right panel shows the SISA bar at 1.4 seconds, dramatically shorter than all other methods at approximately 13 seconds. Bottom-left panel shows SISA accuracy drop near zero, with Label-Flip Retraining approaching the red threshold line. Bottom-right panel shows MIA gap values clustered between 3 and 5 times ten to the negative third.

### 4.5 Cumulative 12-month compliance cost

Figure 3 presents cumulative unlearning time over 12 months (50 requests/month, PATRIC dataset). SISA accumulates 16 s annually versus 160 s for full retraining—a **9.8**× **reduction** in annual compliance burden. On ARMD, the analogous figures are 90 s versus 800 s.

**Figure 3.**
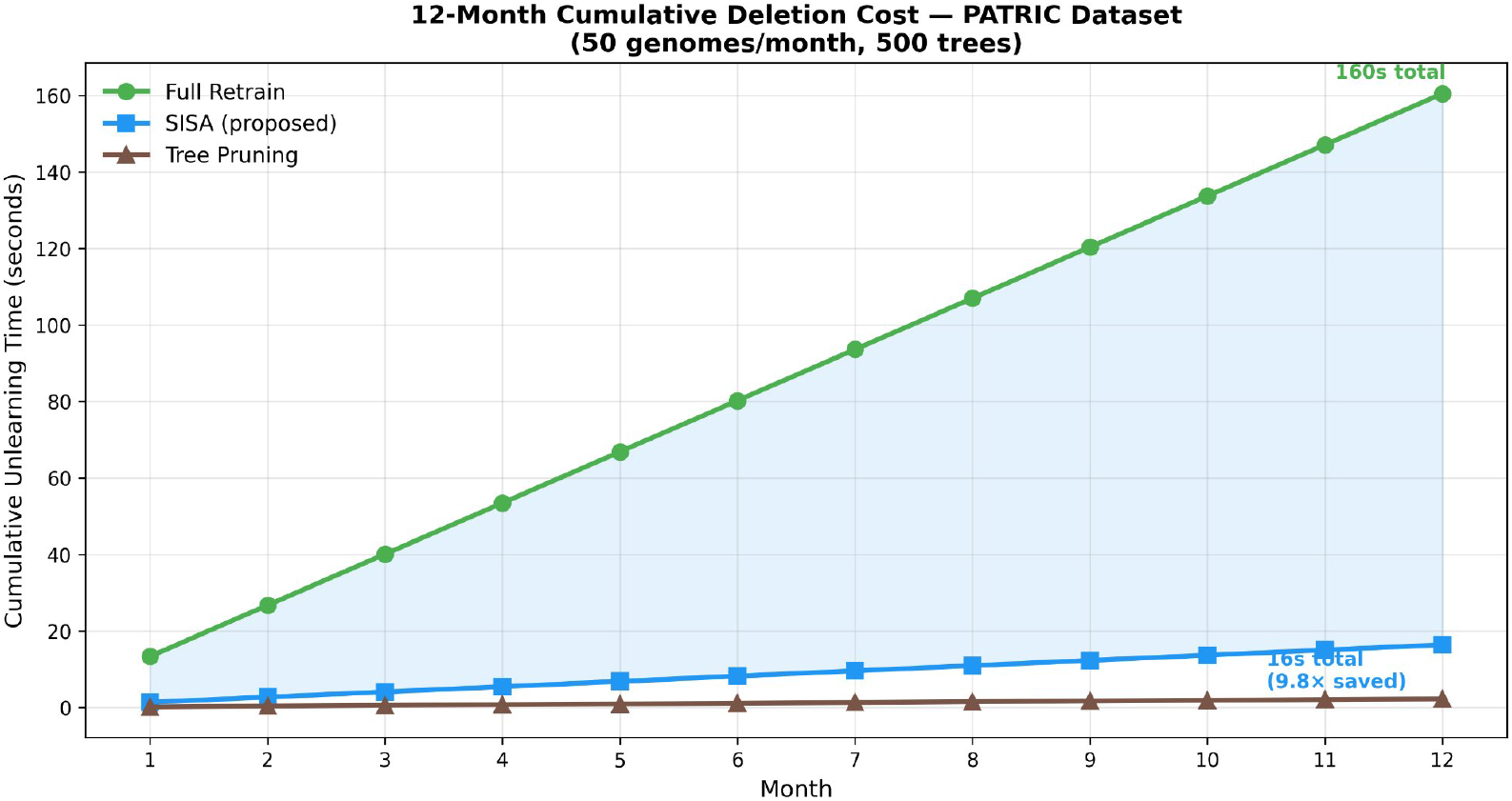
Cumulative unlearning time over 12 months on the PATRIC genomic dataset, assuming 50 deletion requests per month with 500 trees. Full retraining (green circles) accumulates 160 s total compliance overhead versus 16 s for SISA (blue squares)—a 9.8× annual reduction. Tree Pruning (brown triangles) incurs the lowest absolute time (≈2.4 s), but it is excluded from clinical EHR deployment due to the accuracy threshold violations (Table 2). The shaded region highlights the computation time saved by SISA relative to full retraining. Alt text: Line chart showing cumulative unlearning time in seconds over 12 monthly intervals. Full Retrain line rises steeply to 160 seconds. SISA line rises slowly to 16 seconds. Tree Pruning line remains near zero. A shaded region between Full Retrain and SISA represents time savings. Labels at month 12 show 160 seconds total for Full Retrain and 16 seconds for SISA, with a note indicating 9.8 times saved.

### 4.6 Privacy analysis

MIA gaps across all methods and both datasets ranged from 2.591–4.870 × 10^−3^ (ARMD) and 3.226–4.870 × 10^−3^ (PATRIC) a range of ≈ 2.3 × 10^−3^. Notably, the original unmodified model exhibited equally low MIA gaps, indicating that Random Forest ensembles trained on tabular AMR data possess inherent privacy robustness compared to memorisation-prone neural architectures. [16, 21]

## 5. DISCUSSION

Among the five evaluated methods, SISA consistently provided the most favourable balance of computational efficiency and predictive accuracy. By partitioning training data into independent shards, SISA structurally decouples the unlearning cost from the total dataset size, the critical property absent from both Label-Flip Retraining and Influence Reweighting, which require full-dataset training passes and consequently provide no speedup advantage. This is an important negative result for the clinical informatics community: Approximate unlearning methods are not computationally cheaper than full retraining unless they structurally limit the proportion of data retrained.

Tree Pruning’s differential accuracy impact is acceptable on genomic data (−0.045%) but threshold-violating on EHR data (+0.648%) illustrates the risk of deploying modality-agnostic unlearning heuristics without dataset-specific validation. EHR clinical features impose different structural demands on ensemble trees than genomic features; pruning disrupts predictive validity more severely in the clinical EHR setting.

The narrow MIA gap ranges across all methods, including the original model suggests the primary motivation for machine unlearning in Random Forest AMR systems is *regulatory compliance* with GDPR Article 17, rather than mitigation of empirical privacy attacks. This reframing has practical significance: implementation can be governed by regulatory timelines rather than urgent threat response, and SISA’s sub-10-second per-deletion computation is manageable within existing clinical informatics infrastructure. At 7.5 s per unlearning operation, compliance officers can process individual deletion requests interactively, rather than relying on overnight batch retraining jobs with associated model validation and redeployment overhead.

### 5.1 Limitations

The genomic dataset included computationally predicted, alongside experimentally validated phenotypes, which may contribute to lower baseline accuracy (67% vs. 85% on ARMD). The MIA implementation used a confidence-based heuristic; shadow-model attacks may yield different estimates. The experimental setup assumes forget set records concentrate within a single shard, maximising SISA speedup; real-world deletions may span multiple shards and reduce efficiency gains. Only Random Forest classifiers were evaluated; gradient boosting and deep learning AMR predictors warrant separate investigation. Results are reported for a single random seed to ensure deterministic reproducibility; although variance is expected to be small given the large dataset sizes, future work could evaluate robustness across multiple random initializations.

## 6. CONCLUSION

This study presents the first systematic evaluation of machine unlearning methods for AMR prediction models across clinical EHR and genomic data modalities. SISA training achieves 8.9– 9.8× speedup over full retraining with accuracy costs of 0.024–0.048%—below the 0.5% clinical threshold on both datasets and is the only evaluated method satisfying all three requirements for clinical deployment: computational efficiency, accuracy preservation, and cross-modality generalizability. Label-Flip Retraining and Influence Reweighting offer no computational benefit; Tree Pruning is unreliable across data modalities. SISA is readily implementable with standard ML tooling and no specialised hardware and is recommended as the operational standard for GDPR-compliant AMR prediction model maintenance.

## ACKNOWLEDGMENTS

The authors thank the School of Information Technology at Murdoch University for providing computing resources. Artificial intelligence writing assistance tools (including large language models) were used during manuscript preparation; all scientific content, data, analyses, and conclusions are the sole responsibility of the authors.

## AUTHOR CONTRIBUTIONS STATEMENT

### AUTHOR CONTRIBUTIONS

S.and A.A.K. contributed equally to this work.

S.: Conceptualization, literature review, writing original draft, writing review, and editing. A.A.K.: Conceptualization, methodology, software, data curation, formal analysis, investigation, validation, visualization, and writing review and editing.

## DATA AVAILABILITY STATEMENT

The ARMD dataset is available from MIT ClinicalML under a Data Use Agreement at https://clinicalml.org/data/amr-dataset/. The PATRIC/BV-BRC genomic AMR data are publicly available from the NIH Bacterial and Viral Bioinformatics Resource Center via REST API at https://www.bv-brc.org/api/genome_amr/. All experimental code is available from the corresponding author upon reasonable request and is reproducible using the datasets described in the Materials and Methods section.

## SUPPLEMENTARY MATERIAL

Code, preprocessing scripts, and extended result tables are available from the corresponding author upon reasonable request.

